# Characterizing spatiotemporal variation in transmission heterogeneity during the 2022 mpox outbreak in the USA

**DOI:** 10.1101/2023.05.10.23289580

**Authors:** Jay Love, Cormac R. LaPrete, Theresa R. Sheets, George G. Vega Yon, Alun Thomas, Matthew H. Samore, Lindsay T. Keegan, Frederick R. Adler, Rachel B. Slayton, Ian H. Spicknall, Damon J.A. Toth

## Abstract

Understanding how transmission heterogeneity varies over the course of an enduring infectious disease outbreak improves understanding of observed disease dynamics and informs public health strategy. We quantified the spatiotemporal variation in transmission heterogeneity for the 2022 mpox outbreak in the US using the dispersion parameter of the offspring distribution, *k*. Our methods fit negative binomial distributions to transmission chain offspring distributions informed by a large mpox contact tracing dataset. We found that estimates of transmission heterogeneity varied across the outbreak, but overall estimated transmission heterogeneity was low. When testing our methods on simulated data, estimate accuracy depended on contact tracing data accuracy and completeness. Because the actual contact tracing data had high incompleteness, the values of *k* estimated from the empirical data may therefore be artificially high. Through simulation, we explore a method to correct estimated *k* for data incompleteness and, further, explore baseline expectations for temporal dynamics of *k*.

## Introduction

In an infectious disease outbreak, transmission heterogeneity occurs when there is variation in the number of secondary infections per infected host, which can be caused by underlying mechanistic heterogeneity. This variation can manifest at spatial (May and Anderson 1984), temporal (Bjørnstad, Finkenstädt, and Grenfell 2002), and/or individual (Lloyd-Smith et al. 2005; Ke et al. 2022) scales. These different components of transmission heterogeneity interact with one another, creating the potential for complex patterns of transmission heterogeneity over the course of an outbreak. Because transmission heterogeneity plays a critical role in epidemic spread, characterizing how it changes over an outbreak can help to illuminate the causes behind observed epidemic dynamics and inform measures taken by public health decision makers, such as the utility of targeting potentially high-risk individuals or locations for transmission mitigation (Shen et al. 2023; Nielsen et al. 2023).

Prior methods have simultaneously estimated the mean transmission (*R*) and the dispersion (*k*) parameters from data on the number of secondary infections per primary infection (*i.e.*, the offspring distribution) in an outbreak (Lloyd-Smith et al. 2005; Lloyd-Smith 2007; Blumberg and Lloyd-Smith 2013; Toth et al. 2015). These methods have been applied both retrospectively and during ongoing outbreaks (Toth et al. 2016; Kucharski and Althaus 2015; Nishiura et al. 2015) for several diseases. Past research on transmission heterogeneity has largely focused on small outbreaks or has used data from early transmission generations in larger outbreaks (Lloyd-Smith et al. 2005; Lloyd-Smith 2007; Blumberg and Lloyd-Smith 2013; Toth et al. 2015; 2016; Kucharski and Althaus 2015; Nishiura et al. 2015). Some research has explored how the first transmission generation may differ from subsequent generations in *R* and *k* (Toth et al. 2016), but work to understand how individual transmission heterogeneity may vary over the component outbreaks of a large epidemic is more rare (Lau et al. 2020; Tkachenko et al. 2021; Miyama, Jung, and Nishiura 2022).

Among those diseases for which transmission heterogeneity has been characterized in the past is mpox (formerly ‘Monkeypox’ (World Health Organization 2022b)). Monkeypox virus (MPXV), the causative pathogen of mpox disease, is a member of the *Orthopoxvirus* genus of pathogens and is endemic in West and Central Africa. Mpox was first detected in humans in 1970, shortly after the eradication of smallpox (Breman et al. 1980), and has subsequently caused occasional outbreaks thought to result from repeated spillover events (Nalca et al. 2005; Yinka-Ogunleye et al. 2018; Bunge et al. 2022; Durski et al. 2018). While the exact reservoir host is still unknown, the virus has been found in several rodent species native to central and west African countries (Durski et al. 2018). Mpox is known to spread through direct contact with mpox rash, scabs, or bodily fluids; touching objects that have been used by someone with mpox; contact with respiratory secretions; and sexual or intimate contact with an infected person (Centers for Disease Control and Prevention 2022).

Regional outbreaks have been growing in recent years (Sklenovská and Van Ranst 2018; Beer and Bhargavi Rao 2019; Rimoin et al. 2010). Past work to define the basic reproductive number (*R_0_*, the expected number of transmissions from an average infected person in a large, fully susceptible population), for mpox outbreaks have produced estimates below 1, the critical threshold above which an infectious disease will continue to spread and below which an outbreak will eventually die out. However, more recent work has accounted for declining cross-protective immunity from smallpox to reach an estimate of *R_0_* = 2.13 (uncertainty bounds: 1.46-2.67) for mpox in the Democratic Republic of Congo (Grant, Nguyen, and Breban 2020). Past estimates of transmission heterogeneity, quantified by the inversely varying dispersion parameter (low values indicate high transmission heterogeneity) of the offspring distribution (*k*), for mpox outbreaks have been 0.58 (90% confidence interval [CI]: 0.32-3.57) (Lloyd-Smith et al. 2005), 0.36 (0.16-1.47), and 0.33 (0.19-0.64) (Blumberg and Lloyd-Smith 2013), indicating high transmission heterogeneity.

In April 2022, MPXV was identified outside of the countries where the virus is endemic and quickly spread to over 100 countries (World Health Organization 2022a). The World Health Organization (WHO) declared this mpox epidemic a Public Health Emergency of International Concern on July 23, 2022. This rapid global spread was not characteristic of an infectious disease with a basic reproductive number below 1. Contact tracing conducted during the 2022 outbreak indicated that the disease was primarily spreading by sexual contact between gay, bisexual, and other men who have sex with men, often at large gatherings (Philpott et al. 2022). In light of this, public health messaging focused on reducing spread among these groups, especially at large gatherings (Delaney et al. 2022). Potentially due to a novel sexually mediated dominant transmission route, the discrepancy between estimates of the basic reproductive number for mpox outbreaks became globally relevant. Estimates of the 2022 outbreak vary by region and range from 1.56 to 3.8 (Branda, Pierini, and Mazzoli 2022) or potentially higher (Endo et al. 2022).

Because the degree distribution of sexual contact networks can be overdispersed, indicating high variability in the number of contacts per individual, transmission through sexual contact networks can result in heterogeneous patterns of secondary transmission across an infected population (Handcock and Jones 2006). Spreading of the disease through large gatherings, termed ‘superspreader events’, can also cause high levels of transmission heterogeneity (Althouse et al. 2020). Such heterogeneous transmission has been shown to influence outbreak dynamics (Toth et al. 2015; Elie, Selinger, and Alizon 2022) and therefore may have contributed substantially to the rapid global spread of mpox. Modeling studies have provided support for the idea that heterogeneous transmission through contact networks could partially explain the dynamics of the 2022 mpox outbreak (Endo et al. 2022; Spicknall et al. 2022).

Here, we estimate the mean transmission and transmission heterogeneity for the 2022 mpox epidemic in the United States using contact tracing data fortified with algorithmically determined probable case pairs. We estimate these parameters directly from the empirical data and, through simulation, assess the sensitivity of our approach relative to data incompleteness and explore opportunities to correct estimates influenced by data incompleteness. We stratify our analysis by geographic location and time. In this way, we characterize transmission heterogeneity on spatial, temporal, and individual scales. Our aims are to (1) better understand the dynamics of the 2022 mpox outbreak and (2) advance the theory of transmission heterogeneity.

## Methods

### Overview

We used mpox case detection and contact tracing data to algorithmically generate offspring distributions, the number of secondary infections from each case, for given time periods and locations in the United States using methods described in more detail below. Then we used a maximum likelihood approach to fit negative binomial models to those offspring distributions. The negative binomial model fittings provide estimates for the mean *R* and dispersion parameter *k*, with lower values of *k* indicating greater transmission heterogeneity. We also tested our method using synthetic transmission data generated from contact networks known to produce high and low levels of transmission heterogeneity.

### Data

We used data collected through U.S. Centers for Disease Control and Prevention (CDC) mpox case detection and contact tracing in the United States from May 1st to November 7^th^, 2022. This dataset includes, for each probable and confirmed case, information on date of symptom onset, date of reporting, date of positive test, reporting jurisdiction, any recent travel history, and any known contacts with other probable or confirmed cases.

### Algorithmic generation of offspring distributions

To compile an offspring distribution associated with a particular subset of the data, we applied an algorithm to incorporate probable transmission events and their associated infected individuals with unobserved probable transmission events, relying on published mpox serial interval estimates and the dates of symptom onset for infected individuals in the dataset (Figure 1). These transmission trees were constructed using methods similar to those in previous works (e.g., Haydon et al. 2003; Wallinga and Teunis 2004; Heijne et al. 2012), though our method differs in notable ways (see supplemental methods).

**Figure 1.**
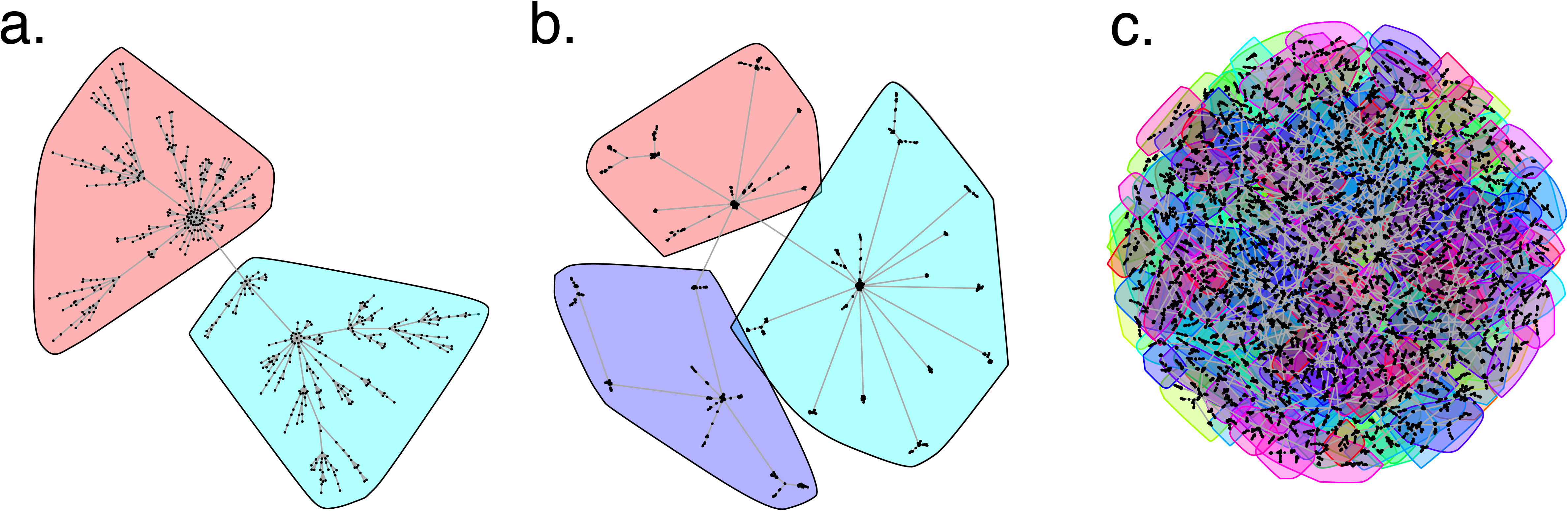
A representation of data processing to build a complete transmission tree. (a) Pre-processed case data points from two public health jurisdictions with four contact tracing identified links. (b) Arrows added to show pools of possible infectors for each case, drawn based on jurisdiction and time (each column of points represents one time step where, for example, possible infectors come from only the most recent two time steps). (c) Chosen infectors sampled and other possible infectors disregarded. This is the post-processing data set used for analysis.

### Parameter estimation

To fit a negative binomial distribution to the offspring distribution, we used a maximum likelihood approach, following established methods (Lloyd-Smith et al. 2005; Toth et al. 2016). This approach is sensitive to the number of individuals with mpox assumed to transmit to no other individuals, which we identify as terminal cases. Because a portion of the individuals in our dataset with no reported offspring may have actually transmitted the disease to others without being detected, including these false terminal cases would artificially inflate the number of zeros in the offspring distribution. To test the sensitivity of our methods to this type of potential bias, we repeated our methods using the zero-inflated negative binomial (see supplemental methods), complementing past work that accounts for underreporting of terminal cases (Lloyd-Smith 2007).

In the negative binomial estimates, the logarithms of negative binomially distributed probabilities of each data point are a function of *R* and *k*, while in the zero-inflated estimates the logarithms of zero-inflated negative binomially distributed probabilities of each data point are a function of *R*, *k*, and *p_0_*. We used maximum likelihood estimation by taking the log-likelihood function of the entire data set. We then used multivariate optimization to find *R* and *k* for the negative binomial estimates and *R*, *k*, and *p_0_* for the zero-inflated estimates. We used the likelihood ratio test to find 95% confidence intervals for these parameters.

### Sliding and telescoping approaches

To evaluate change in parameter estimates over time, we conducted our fitting methods on offspring distributions generated across both sliding (fixed width) and telescoping (varying width) windows of time (see supplemental methods).

### Simulations

To characterize the reliability of our methods to estimate transmission heterogeneity, we repeated them on simulated data generated with a model of infectious disease spread over a static contact network. We conducted simulations on undirected scale-free networks, shown to produce heterogeneous transmission (Bansal, Grenfell, and Meyers 2007). We initialized the scale-free network with a Barabási-Albert algorithm of preferential attachment (Barabási and Albert 1999), allowing for the network to serve as a proxy for a population with highly heterogeneous contact rates where simulated outbreaks can be expected to result in an overdispersed offspring distribution. The network was generated using the *barabasi.game* function from the *igraph* R package (Csardi and Nepusz 2006), using a population of 50,000 and power of 1.

Each outbreak was seeded with a randomly chosen single infection. Disease dynamics were modeled following a *Susceptible – Exposed – Infectious – Recovered* paradigm, parameterized for mpox. The latent period was a random variable drawn from a lognormal distribution with log mean = 1.5 and log standard deviation = 0.7 (Madewell et al. 2022). For lack of detailed information on the duration of infectiousness for mpox, the infectious period in our simulations was a random variable drawn from a gamma distribution with shape = 21 days and scale = 1. This distribution is consistent with the reported serial interval distribution (Madewell et al. 2022), clinical guidance indicating symptoms last 2-4 weeks (Word Health Organization 2022c), and investigations reporting that some hospitalized cases show symptoms for much longer periods (Adler et al. 2022). However, it should be noted that more detailed study may reveal that the distribution we used may not describe the infectious period for mpox. Contacts of infectious nodes were infected with probability of 0.15 per contact day.

In order to replicate our methods on the simulated data, we assigned each node a “jurisdiction” using a modularity optimization algorithm that identifies community structure in networks (Blondel et al. 2008), as depicted in Figure 2. We then identified the most probable infector for each case, as detailed above, and estimated the parameters *R* and *k* by fitting a negative binomial distribution, as detailed above.

**Figure 2.**
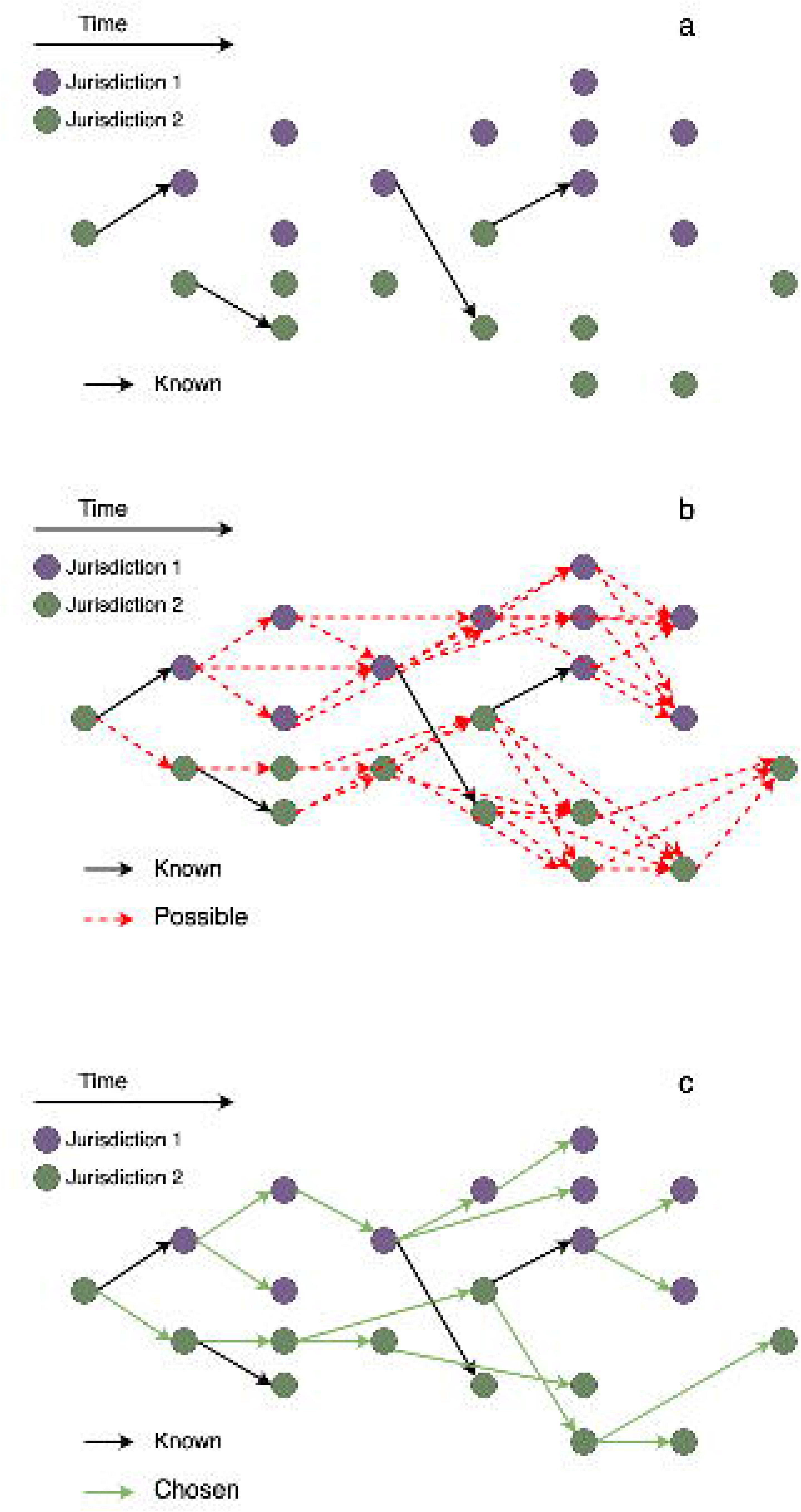
Visualizations of the contact network used in simulations, with simulated jurisdictions shown in colored polygons. See text for description of methods used to generate networks; (a) shows two jurisdictions, (b) shows three jurisdictions, inclusive of the two in (a), and (c) shows the full contact network. Overlapping polygons in (c) are visual artifacts of the large network –each node is associated with only one jurisdiction.

In order to test the sensitivity of our methods to incompleteness in detailed contact information, we repeated the methods with different levels of incompleteness. Proportions of data completeness (*f*) ranged from 0 to 1, and we explored two types of incompleteness. For the first type of incompleteness (“missing nodes”), individuals for which detailed contact information was completely excluded were randomly sampled from the simulated population with equal probability. For the second type of incompleteness (“missing edges”), connections between individuals to be excluded from the dataset were randomly sampled with equal probability. Methods were repeated on each level of completeness, and the resulting *k* estimates at the end of the outbreak using a telescoping window were compared. The relationship between *f* and *k* was described by fitting linear and exponential model to the data (log transforming the response variable for the exponential model) using the ‘lm’ function and goodness-of-fit was assessed with the Akaike Information Criterion using the ‘AIC’ function in R statistical software (R Core Team 2017).

To evaluate the effectiveness of our method in estimating *k* from data on outbreaks where high transmission heterogeneity is not expected, we simulated an outbreak over a version of the contact network used in the first simulation modified by connecting all nodes within jurisdiction clusters with edges and maintaining the existing cross-jurisdiction edges. The modified network therefore has very high connectivity within jurisdiction clusters (i.e., jurisdiction subgraphs are complete, representing very well mixed local populations), but connectivity across jurisdiction clusters is equal to that of the unmodified network. We repeated our methods as detailed above, estimating parameters *R* and *k* across different levels of incompleteness in detailed contact information.

All computational methods were conducted in R statistical software (R Core Team 2017).

## Results

### National estimates

There were 22,391 probable and confirmed mpox cases in the empirical dataset, after excluding 5,074 (18.47%) cases with missing data on symptom onset date. In the exclusive dataset, 820 (3.66%) of these cases had contact tracing-identified connections to other probable and confirmed cases, compared to 12 (0.24%) of those excluded cases.

*R* and *k* estimated at the national level changed over time. Using a sliding 2-week window (Fig. 3a), we observe that, after early increase to a peak, estimated *R* declines over the course of the epidemic, dropping below 1 around the peak of new infections on approximately August 1st, as is expected to occur when an outbreak changes from growth to decline. Median estimated *k* remains above 1 for all time windows, though with high variability – a pattern which does not indicate high transmission heterogeneity.

**Figure 3.**
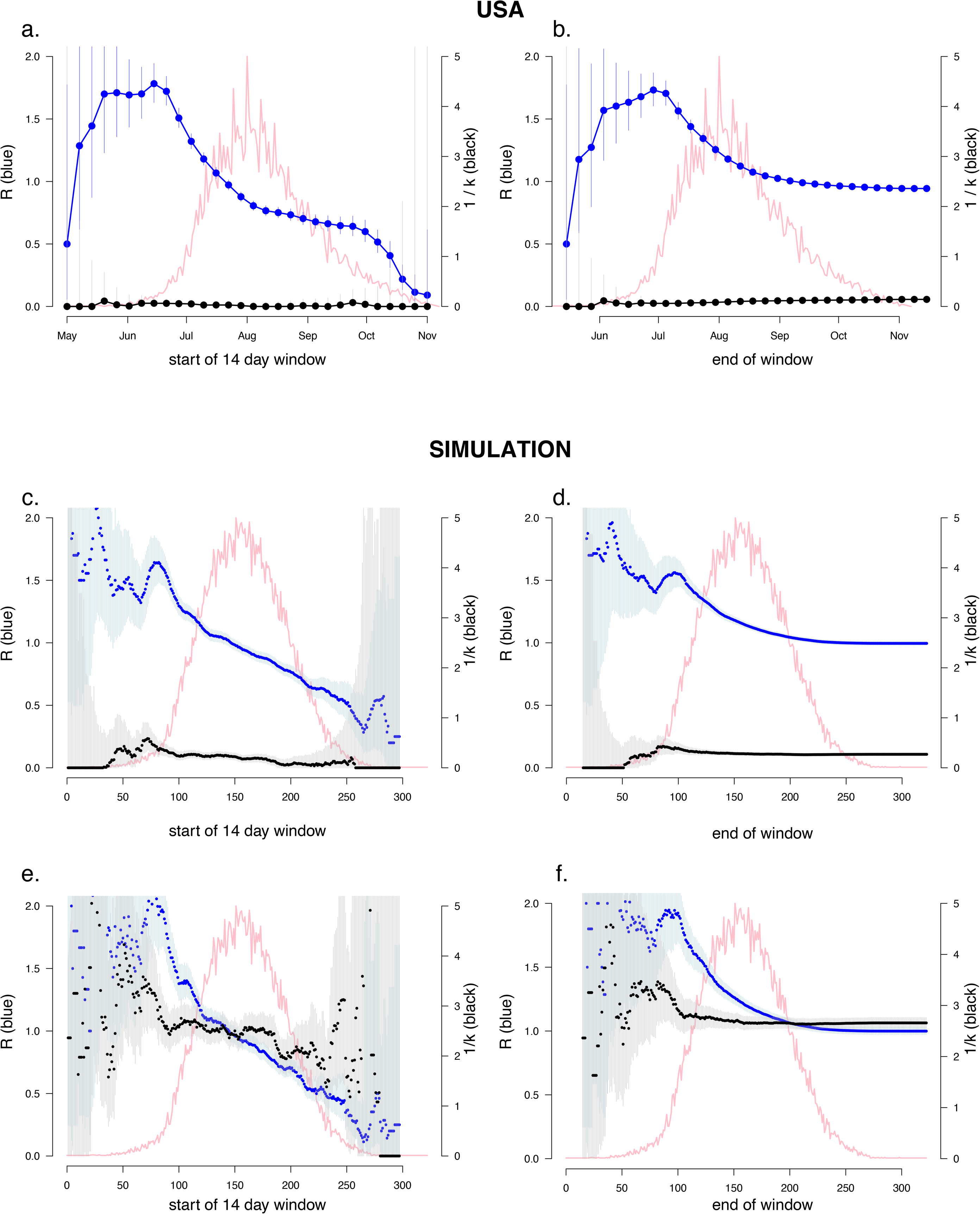
Temporal variation in transmission parameters. Left column shows estimates made with a sliding window, right column shows estimates made with a telescoping window, starting on May 1st, 2022. Time points for the sliding window approach (left column) are the start of the 14-day sliding windows, time points for the telescoping window approach (right column) are the end of the telescoping windows. Top row shows empirical parameter estimates, middle and bottom rows show estimates made from outbreaks simulated on a scale-free contact network. In the middle and bottom rows, the horizontal axis is units of days since the start of the outbreak simulation. In all plots, black points represent values of 1/k, plotted on the right vertical axes, and blue points represent values of R, plotted on the left vertical axes. (a-b) Estimated median and 95% CIs for 1/k (black, right-hand vertical axis) and R (blue, left-hand vertical axis), displayed over daily new mpox cases (pink, peak of cases in empirical dataset in panels a,b is 506 on August 1^st^, 2022) for the 2022 mpox epidemic in the United States. (c-f) Parameter values estimated on simulated data from a simulated outbreak which exhibited strongly heterogeneous transmission. Peak of cases (pink) in simulated dataset is 410 on day 151 of the simulation. (c-d) Estimates from simulated data where probable transmission pairs were determined with complete information on true individual contacts for 3.6% of the population, with the remainder being determined only with simulated dates and geographic locations. This approximates the condition of the empirical data in a-b. (e-f) Estimates from simulated data where all pairs were determined with complete information on true individual contacts. In all plots, values of 1/k, instead of k, are displayed to ease visualization.

Applying a telescoping window approach allows us to evaluate *R* and *k* taken cumulatively at different points during the epidemic (Fig 3b). Using the time window beginning at the start of the outbreak, May 1^st^, and ending at the peak of new cases, August 1^st^, we find that *R* = 1.25 (95% CI: 1.21-1.28) and *k* = 12.18 (95% CI: 8.18-21.27). Using a zero-inflated negative binomial distribution returned the same values of *R* and *k* for this period, with a *p*_0_ value of 0.001 (95% CI: 0.00-0.02).

### Jurisdiction estimates

Estimates of *R* varied across U.S. State and over time (Fig. 4). Estimates of *k* also vary across public health jurisdictions and over time (Table S1). While median estimates of *k* are over 1 at all time points for every jurisdiction, the variation in *k* estimates across iterations is substantial for some points, indicating a greater potential for higher transmission heterogeneity at that time and place. Estimates of *p_0_* made using the zero-inflated negative binomial were highly variable across jurisdictions and timepoints (see supplemental results).

**Figure 4:**
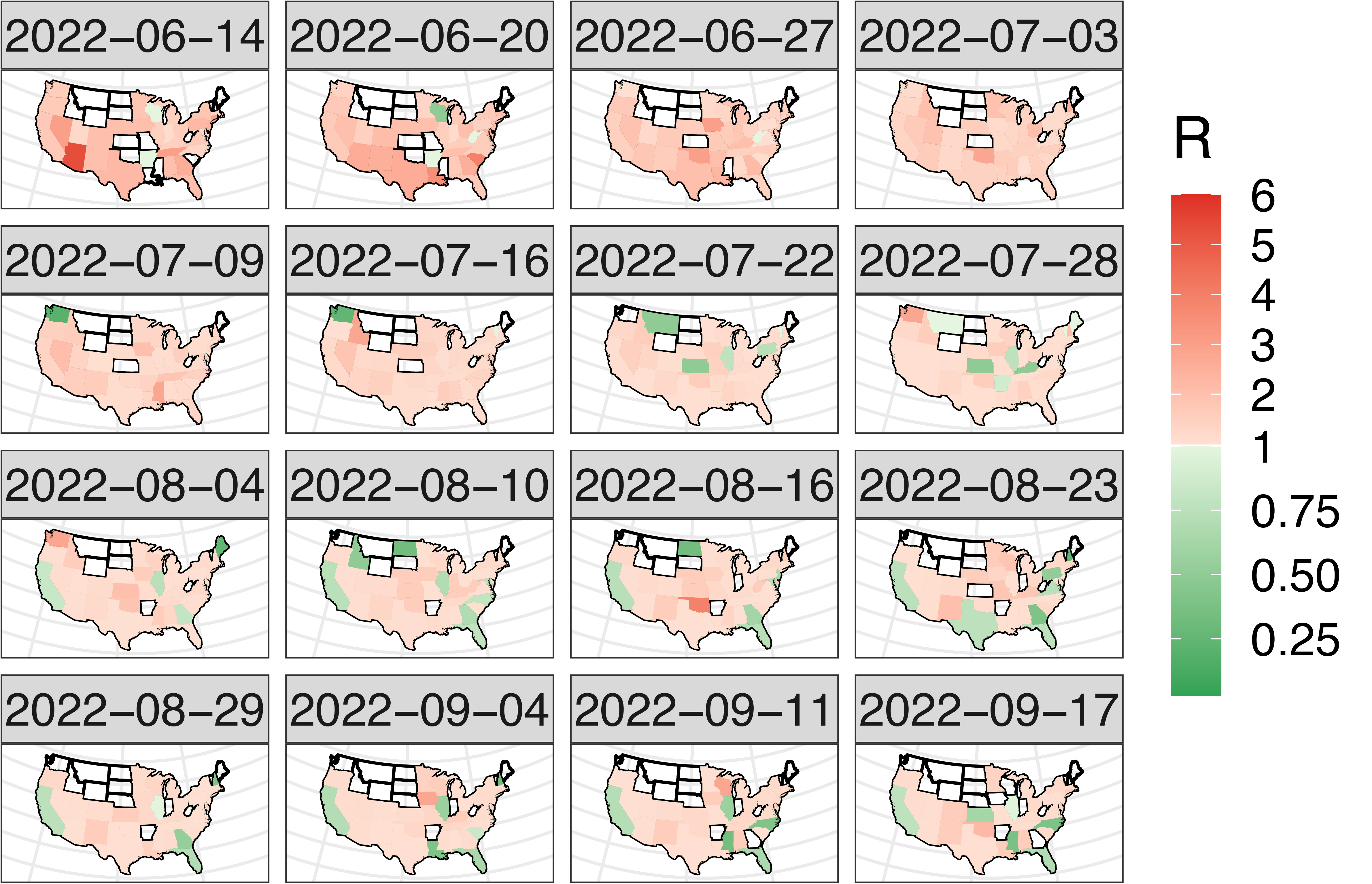
Spatiotemporal variation in estimated values of R across US states during the middle of the 2022 mpox epidemic. Unshaded regions indicate insufficient data to conduct parameter estimations for that region and time period.

### Simulations

Conducting the estimation methods on data resulting from an outbreak simulation over a scale-free contact network and with a gamma-distributed infectious period indicated heterogeneous transmission, as anticipated, with *k* estimates well below 1 with complete data (Fig 3e,f; Fig 5a-d). Time-varying *k* estimates acquired through the use of a sliding window were low at the start and increased over the course of the outbreak. Conducting the methods on data from an outbreak over the modified network showed that transmission was less heterogeneous overall, but time-varying *k* estimates showed pulses of values less than 1, indicating temporary periods of high transmission heterogeneity after which heterogeneity declines (Fig 5e,f).

**Figure 5:**
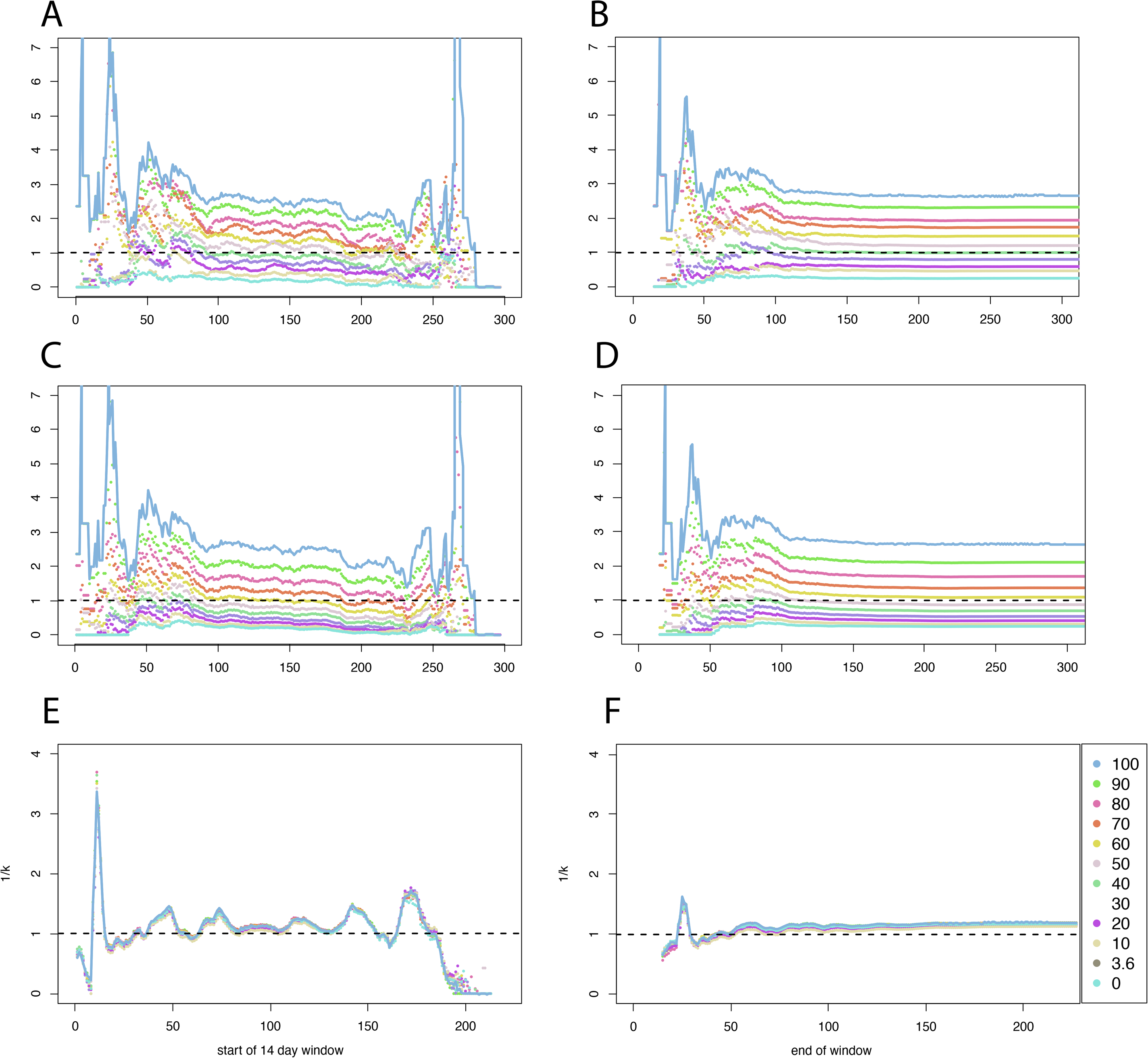
Estimates of k over time from a simulated outbreak over a scale-free (a-d) and a modified contact network with high within-jurisdiction connectivity (e,f), using sliding (left column) and telescoping (right column) analysis windows. Panels a-b show “missing node” mode of missingness, panels c-d show “missing edge” mode.

The method showed reliability in detecting overdispersed offspring distributions when detailed contact information was abundant, reflected by estimates of *k* below 1 when *f* was high (Fig. 3e,f). However, under the missing node paradigm with *f* = 0.036 (i.e., when data missingness was equivalent to the estimated missingness in the empirical dataset), estimates of *k* were much higher, staying above 1 (Fig. 3c,d). By varying the proportion of the simulated population for which detailed contact information was used, we explored the relationship between *k* estimate accuracy and data completeness (*f*). This relationship is approximately linear (AIC: –19.3 versus –4.59 for best-fit exponential model) and can be described by the equation

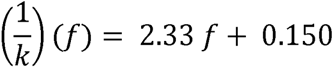

where *1/k* is the value of *1/k* at the end of the simulation using a telescoping window, and *f* is the fraction of data that is complete (Fig. 6a). Under the missing edge paradigm with *f* = 0.036, estimates of *k* were also above 1 (Fig. 6c). Varying *f* under this paradigm, we recover an exponential relationship (AIC = –39.2 versus 2.23 for best-fit linear model) between *k* and *f* as:

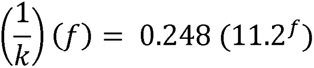

**Figure 6.**
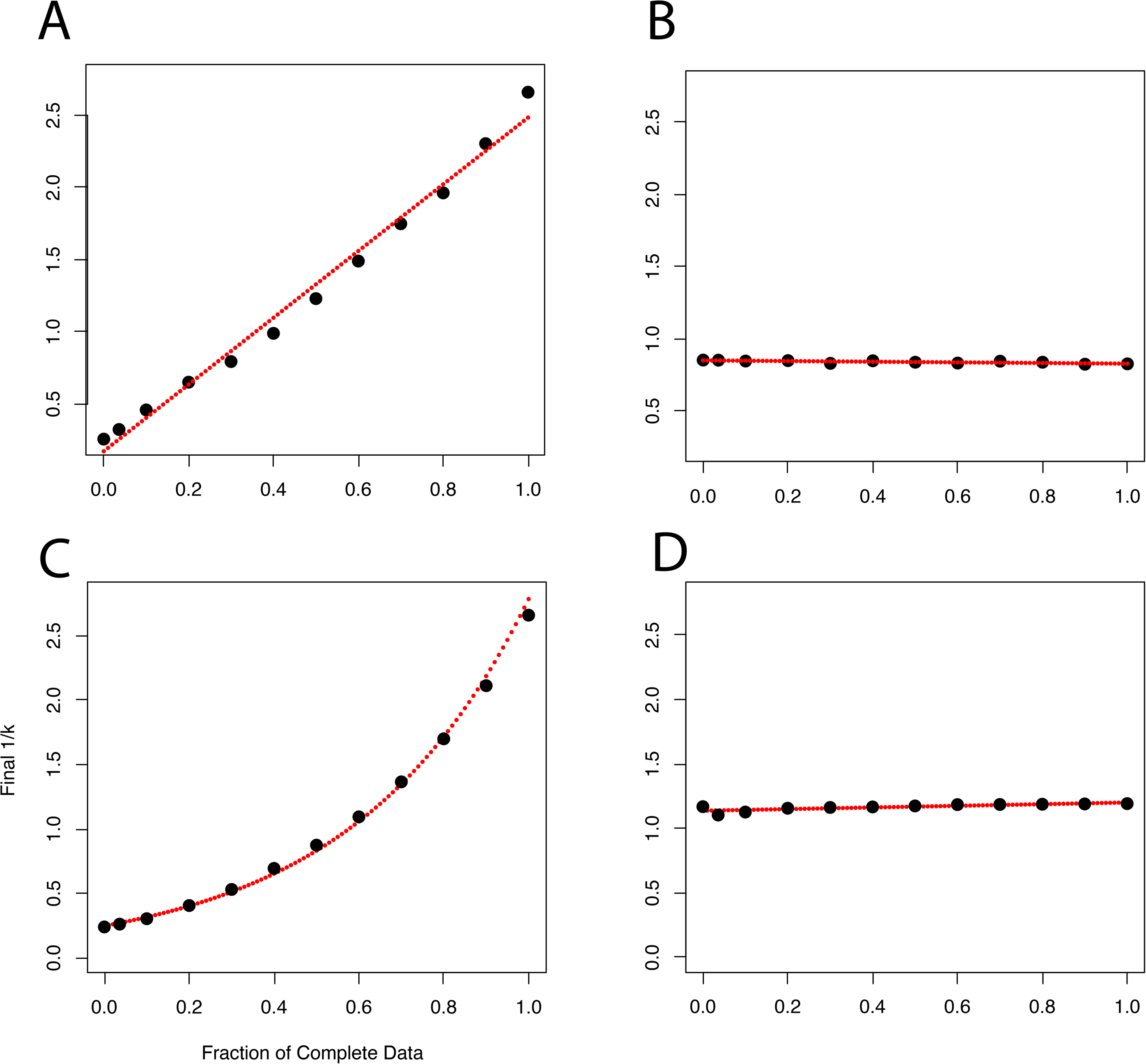
Estimates of 1/k on highly heterogeneous outbreaks over a scale-free contact network (a,c) and low heterogeneous outbreaks over a modified contact network (b,d), at the end of a simulation using methods applied to various fractions of complete detailed contact information plotted as black points, with red dotted line describing best-fit models (1/k)(f). Panels a-b show “missing node” mode of missingness, panels c-d show “missing edge” mode.

When the methods to estimate *k* were conducted on data from an outbreak simulated over a modified contact network with high within-jurisdiction connectivity (see methods), estimates of *k* were around 1 (Fig. 5e). *k* = 1 is a special condition of the negative binomial: the geometric distribution. A geometric offspring distribution is expected under the assumption of constant infectiousness and a constant recovery rate across infected individuals in a well-mixed population (Grassly and Fraser 2008). At times, estimates of *k* drop well below 1 (observed as spikes in 1/*k* in Fig. 5e). These instances may occur simultaneously with pathogen invasion of a previously uninfected jurisdiction cluster, and the transient jump in heterogeneity may reflect a sudden increase in the effective susceptible population followed quickly by a sharp decline in the same. Estimates of *k* acquired with a telescoping window approach tended towards 0.83 by the end of the outbreak (Fig. 5f), reflecting the cumulative influence of these temporary spikes in transmission heterogeneity over the course of the outbreak. In contrast to the results from the scale-free network-based simulation (Fig. 5a-d), *k* estimates are very near their true values regardless of the degree of data incompleteness (Fig. 5e,f). Fitting a linear model produces equation

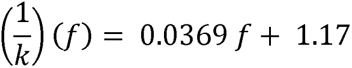

with the small slope indicating constancy (AIC = –82.2 versus –77.9 for exponential).

### Correcting empirical estimates to account for data incompleteness

Ideally, the empirical dataset in this study would be sufficient to derive a correction function, as with the simulated data, above. However, with only 3.6% completeness and substantial noise (discussed below), a model fit to a set of *k* values estimated from datasets with artificially increasing incompleteness may be unreliable. To explore possible values for corrected *k* estimates, we proceeded to use a correction factor derived from simulated data on our estimates from empirical data.

Under the assumption that the 2022 mpox outbreak in the United States occurred over a contact network that was roughly scale-free (i.e., where the degree distribution follows a power law) (Murayama et al. 2023), the function derived from our simulation results can be used to correct our estimates from empirical data to account for data incompleteness. Doing so yields much lower estimates of *k*. Using the slope of the linear model from the high heterogeneity simulations under the missing node paradigm, we can use our data point (*f* = 0.036, *k* = 7.06) to find equation

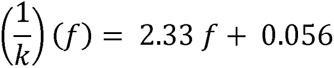

Therefore, when *f* = 1 (i.e., under complete detailed contact information), *k* = 0.418. This value of *k* indicates high heterogeneity and is in line with past estimates of *k* for mpox (Lloyd-Smith et al. 2005; Blumberg and Lloyd-Smith 2013). The validity of this corrected estimate is highly contingent upon the assumption that the contact network properties of the real mpox outbreak are similar to the simulated network. While there is some evidence that this could be the case (Murayama et al. 2023), care must be taken with any applied interpretation. If the contact network of the real outbreak was, instead, evenly mixing within jurisdictions (as in simulations over a modified contact network, Fig. 5e,f), correcting for incompleteness would impact *k* estimates far less, and the corrected *k* estimate may not indicate high heterogeneity.

## Discussion

Our estimates of transmission heterogeneity over the course of the 2022 mpox outbreak in the United States were low, but these estimates may be biased due to data incompleteness. If taken at face value, our estimates of *k* garnered from direct analysis of the real data suggest that transmission of mpox during the 2022 outbreak in the United States was not highly heterogeneous; estimates of *k* >1 across many jurisdictions and time periods would seem to indicate that transmission was approximately what would be expected in an evenly mixing population of people with equal inherent transmissibility, if not lower. We do, however, find some indication of overdispersed transmission in certain locations during certain time periods (Table S1), which could be consistent with explanations that invoke individual level transmission heterogeneity.

However, if the context of data incompleteness is taken into account and it is assumed that contact network structure was roughly scale-free, corrected estimates of *k* could, under certain conditions of data missingness, indicate that transmission of mpox was highly heterogeneous. This second possible conclusion hinges heavily upon the assumption that the contact network of the real mpox outbreak was roughly scale-free. If it is not, then the function to correct *k* based on data missingness would have a low slope, the effect of data incompleteness on *k* estimates may be much smaller (as in fig 5e,f), and therefore the empirical estimate could be seen as accurate.

The contact network structure and the data quality in our empirical dataset likely varied by geographic location and through time. Therefore, we cannot rule out the possibility that our estimates of *k*, corrected or otherwise, for the 2022 US mpox epidemic are artificially high in some cases while artificially low in others, and care must be taken with any interpretation. Still, our procedure to correct for data incompleteness under the assumption of a proposed contact network structure leads our *k* estimates to be more in line with past estimates of transmission heterogeneity for mpox (e.g., Lloyd-Smith et al. 2005; Blumberg and Lloyd-Smith 2013).

Our study contributes to a small but growing literature on spatiotemporal variation in transmission heterogeneity of human infectious diseases. Past research on transmission heterogeneity has largely focused on small outbreaks or has used data from the first generations of infections in larger outbreaks. Related research on the influence of contact networks on the spread of infectious diseases has illuminated important concepts regarding transmission heterogeneity. For example, as the variation in the degree distribution of the infected population changes over the course of an epidemic simulated on a network with a particular underlying structure (Bansal, Grenfell, and Meyers 2007), transmission heterogeneity should be expected to change over time. Our study provides a description of the baseline expected temporal change in transmission heterogeneity for outbreaks on scale-free contact networks with non-constant infectious period: generally, transmission heterogeneity reaches an early peak and then declines over the course of an outbreak.

These methods are adaptable to other infectious disease outbreaks. They could provide utility in informing public health decision makers about time-varying transmission heterogeneity, with a lag determined largely by the time from infection to reporting. The method we present to correct for incompleteness is robust to variation in transmission mode and contact network structure across outbreaks, given contact information for a sufficiently large proportion of the population. The degree of incompleteness in the empirical dataset in the present study did not reach a threshold that allowed us to derive a reliable correction function from that data, so we used our simulated data, parameterized for mpox, as a proxy to derive a correction function to apply to the empirical data. Ultimately, this data limitation reduces our confidence in our corrected estimate, and care must be taken with interpretation. Future work to apply the method we present here to ongoing infectious disease outbreaks will require solutions to these data limitations.

## Supporting information

supplemental discussion

## Data Availability

Code to replicate the simulation-based methods will be made publicly available upon acceptance for publication. Data used in the empirical analysis is individually identifiable and will not be made publicly available.

## Acknowledgements

The findings and conclusions in this report are those of the authors and do not necessarily represent the views of the Centers for Disease Control and Prevention. JL, TRS, GGVY, AT, MHS, LTK, and DJT acknowledge research support by the Centers for Disease Control and Prevention (1U01CK000585; 75D30121F00003)

## Notes

### Author Declarations

This activity was reviewed by CDC and conducted consistent with applicable federal law and CDC policy.

### Summary of Updates

Manuscript updated in response to peer review.

