## supplemental discussion for "Characterizing spatiotemporal variation in transmission heterogeneity during the 2022 mpox outbreak in the USA"

Incomplete contact tracing data can be the result of various errors introduced during the data collection process. Our methods operated under a number of assumptions related to missing data. First, it was assumed that the data are a representative sample of all mpox cases. If there is an association between case detection and secondary infections produced, then this assumption could impact our estimates. For example, diagnostic testing can be biased towards cases of more severe disease (Tsang et al. 2021), which could produce more secondary infections through an increased duration of viral shedding (Fielding et al. 2014). Second, we excluded the cases lacking symptom onset dates, and we assumed that the cases for which date of symptom onset exist constitute a random sample from the population. If there is a relationship between having the symptom onset date for a case and some other factor (e.g., information on contacts), then our results could be impacted.

To account for possible undercounting of secondary transmissions, we repeated our estimates by fitting zero-inflated negative binomial distributions. Undercounting of secondary transmissions may manifest as extra zeros in the offspring distribution, which can be introduced in the following three ways. First, inconsistencies in the dates associated with cases could impact our analyses. Date of reporting could occur at any time during the course of an infection, even after the patient is no longer infectious, and could be influenced by “data dumps”, where particular jurisdictions may have posted large portions of their data on a given day. While we use date of symptom onset, it is possible that these dates were influenced by behavioral or systematic processes in healthcare and disease progression. Even for cases with a date of symptom onset, we cannot be sure those dates are consistent with the start of the infectious period. Second, there are barriers to a suspected case becoming a probable or confirmed case – tests may only be provided to verified contacts of another probable or confirmed case or to those in high-risk subpopulations. In the extreme, this feature of the contact tracing process can result in missing entire transmission chains. Third, identifying case-by-case contacts and their exact dates can be difficult in sexual/social networks such as those where mpox has been shown to spread. Infected individuals may not know or choose to disclose the identities of their close contacts and additionally may delay seeking care due to historical discrimination and stigmatization of the LGBTQ+ community in the healthcare setting (Ennab et al. 2022; Rodríguez et al. 2022; The Lancet Regional Health – Europe 2022). The spatiotemporal variability in estimates of *p*_0_ may reflect variation in data incompleteness related to this potential zero inflation. However, overall at the national level, estimates of *p*_0_ were consistently low, and our sensitivity analysis using the zero-inflated negative binomial distribution returned very similar parameter estimates to those returned using the negative binomial distribution. In a large population, the influence of missing case data on estimates of *R* and *k* may be minimized or obscured.

One reason that transmission heterogeneity is of potential interest to public health decision making is that it can indicate that within-host processes during infection result in inherent differences in the probability of infection given contact across infected people (Ke et al. 2022). In such a situation, equal rates of contact between infected and susceptible people will result in different numbers of secondary infections, and interventions, when implemented, may differ in their efficacy relative to expectations. Additionally, we should expect to find consistently low values of *k* across spatiotemporal dimensions. In the case of the 2022 mpox outbreak in the United States, we did not find direct evidence that this sort of individual-level transmission heterogeneity was broadly present. Setting aside the corrected *k* estimates, the rare occurrences of highly heterogeneous transmission that we observe could have resulted from differences in contact rates, either due to standing individual variation in contact rates or to the occurrence of superspreading events. In this event, targeted interventions could be employed to reduce the spread of disease. Without high heterogeneity in transmission, the rate of disease spread is well-characterized by an *R*_0_ value greater than the threshold 1, as has been suggested by other recent studies of *R*_0_ for the 2022 mpox epidemic (Branda, Pierini, and Mazzoli 2022; Endo et al. 2022). These results are consistent with the theory that cross-protective immunity from smallpox may be waning, resulting in higher rates of overall transmission (Grant, Nguyen, and Breban 2020).

It is important to recognize that our corrected estimates of *k* under the “missing nodes” assumption are consistent with past estimates for mpox. However, since these corrected values are contingent upon an assumption of contact network structure, a discussion of the disparity between our uncorrected estimates (*k* >1) and those of past work which estimated low values of *k* for mpox outbreaks is justified. The population structure, timing, environmental characteristics, and primary mechanisms of transmission differed between the 2022 mpox outbreak in the United States and the outbreaks described in other studies. It is possible that these differences explain the differences in estimates of *k* between our study and the others. The timing of the US outbreak relative to the beginning of the global epidemic may also have afforded individuals with a higher potential risk of infection the time necessary to take precautions against infection. This may be especially true as it relates to large events, which were implicated in the initial spread outside the United States and were the focus of targeted public health messaging in the United States (Delaney et al. 2022). Indeed, survey results suggest that many US gay and bisexual men and nonbinary and transgender people took active measures to reduce the risk of exposure to mpox (Delaney et al. 2022), and so this could explain the values of *k* we observed.

Molecular, phylogenetic, and in-vivo studies have indicated that monkeypox virus samples from the outbreak in this study are significantly divergent from those of past outbreaks (Isidro et al. 2022, Americo et al. 2023). The outbreak investigated in the current study is caused by a virus that might have quite different epidemiological characteristics than the virus associated with outbreaks upon which past estimates of epidemiological parameters have been based. Our methods can be extended to clarify the differences between our estimates of *k* and those of past studies of the same disease.

Our methods used a straightforward algorithm to determine likely infectors for cases without a contact tracing-identified infector, and we subjected these methods to a limited sensitivity analysis using simulated data. In the process, we determined that our methods can be expected to accurately estimate *k* with high-quality data on the direct contacts of each case. In some instances, our empirical data may meet these qualifications, but in other instances they likely do not. So, it is possible for our methods to mask high transmission heterogeneity under some conditions. We provide a framework to correct estimates of *k* for data incompleteness, but extensions to this framework would be necessary to provide robust, reliable estimates in a public health setting. One study has found that different models used to measure transmission heterogeneity may result in different estimates with starkly different interpretations, with their instant-individual heterogeneity model recovering high heterogeneity when alternate models do not (Zhang, Britton, and Zhou 2022). The methods we present here are adaptable to incidence time series data, holding promise that transmission heterogeneity may be reliably and easily estimated, but care must be taken to contextualize findings within the caveats of the data and the potential limitations of data gathered during emerging outbreaks.
