## supplemental methods for "Characterizing spatiotemporal variation in transmission heterogeneity during the 2022 mpox outbreak in the USA"

*Processing*

To compile the offspring distribution, we started by including contact-tracing identified probable transmission events and their associated infected individuals. For each infected individual for whom a probable infector has not been identified by contact tracing, we used dates of symptom onset included in the dataset and published serial interval estimates to compile a pool of possible infectors (Fig. 1). We defined a possible infector of an infected individual as one with overlapping location (US county) and whose date of symptom onset is within one maximum serial interval, 21 days, of the other’s date of symptom onset (UK Health Security Agency 2022). Because the pool of possible infectors generally includes more than one possible infector, we then sampled one possible infector from each pool to serve as the chosen infector for each infected individual. In this sampling process, the serial interval was calculated for each possible transmission, and the probability that a serial interval was sampled is equal to the gamma distribution density, parameterized according to published estimates of the serial interval distribution for mpox (gamma distribution, shape = 2.9, scale = 2.9; Madewell et al. 2022). The individual producing the sampled serial interval was assigned to be the infector for an infected individual. From this dataset, containing both contact-tracing identified and algorithmically determined transmission pairs, we calculated the number of secondary infections produced for each infected individual and fit a negative binomial distribution to the resulting offspring distribution, finding the best-fit parameter values and their 95% confidence intervals. This entire process was repeated 20 times with replacement, producing a sample of possible offspring distributions of an outbreak

*Sliding and telescoping approaches*

To evaluate change in parameter estimates over time, we conducted our fitting methods on offspring distributions generated across both sliding (fixed width) and telescoping (varying width) windows of time. In the sliding window approach, offspring distributions were generated consecutively over 14-day periods, with each consecutive window starting 7 days after the start of the previous window (a 50% overlap). The sliding window approach generates estimates that can be likened to parameters *R_t_* and *k_t_* described elsewhere (Schneckenreither et al. 2022; Zhang, Britton, and Zhou 2022; Wallinga and Teunis 2004), though to avoid confusion regarding definitions and assumptions that differ from those of our estimated parameters, we simply use *R* and *k*. In the telescoping window approach, consecutive windows are inclusive of the previous window; the end date for each consecutive window is advanced 7 days while the start date remains the same, May 1st, 2022. Estimates of *R* and *k* generated from the telescoping window approach describe the mean and dispersion of secondary transmission from the beginning of the outbreak up to the end date of a given time window. In this way, they can be thought of as the effective *R* and *k* parameters, but again we simply use our previously defined *R* and *k* terms for the mean and dispersion of the offspring distribution.

*Parameter estimation*

The zero-inflated negative binomial differs from the negative binomial distribution in its inclusion of a parameter *p_0_*, the probability that an individual’s onward transmission data are absent, independent of the number of individuals they infected, leaving 1 - *p_0_* as the probability that the individual’s transmission data are complete. A lower *p_0_* indicates greater confidence in the accuracy of the data, and the zero-inflated negative binomial reduces to the negative binomial when *p_0_* = 0. Similar to past work that fits modified negative binomial distributions to discrete data that undercounts zeros (Ridout, Demetrio, and Hinde 1998; Lloyd-Smith 2007), accounting for *p_0_* helps to correct for missingness in the data and make more reliable estimates of *R* and *k*. The probability mass function (pmf) for the zero-inflated negative binomial can be defined as follows, with the reported number of offspring for each infected individual distributed based on whether or not the data point, *y*, is a zero, where we assume the true number of offspring is negative binomially distributed:

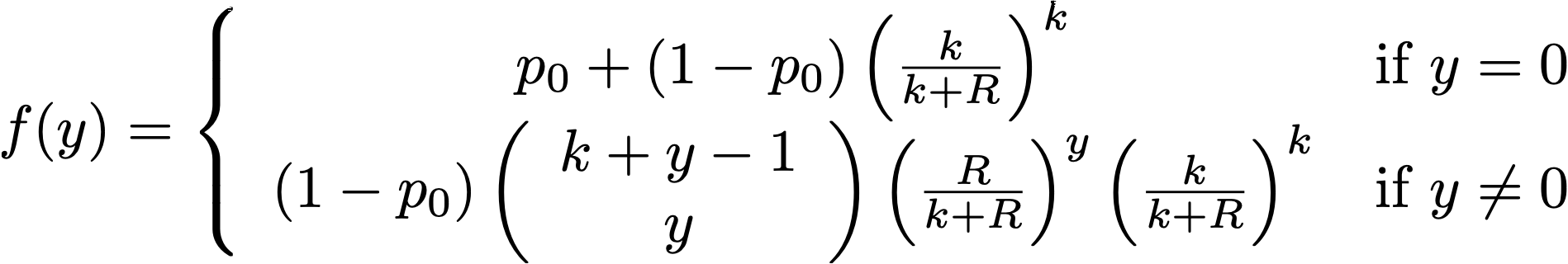
